# Breadth of neutralising antibody responses to SARS-CoV-2 variants of concern is augmented by vaccination following prior infection: studies in UK healthcare workers and immunodeficient patients

**DOI:** 10.1101/2021.06.03.21257901

**Authors:** Angalee Nadesalingam, Diego Cantoni, David A Wells, Ernest T Aguinam, Matteo Ferrari, Peter Smith, Andrew Chan, George Carnell, Luis Ohlendorf, Sebastian Einhauser, Ralf Wagner, Nigel Temperton, Javier Castillo-Olivares, Helen Baxendale, Jonathan L Heeney, the HICC consortium

## Abstract

Approximately 75% of the UK population has received only one dose of a 2-dose COVID-19 vaccine regime in the face of circulating SARS-CoV-2 Variants of Concern (VOCs). We aimed to determine the levels of vaccine-induced neutralising antibodies to SARS-CoV-2 variants B.1.1.7, B.1.351 and P.1. To do so, we undertook a single-centre cross-sectional study of health care workers (HCWs) and outpatients with immunodeficiencies (IDP) based at the same critical care tertiary NHS Trust, following a single dose of either BNT162b2 or AZD1222 vaccines. Data revealed low neutralising antibodies (nAbs) in IDPs, with only 5% and 3% showing detectable neutralisation of B.1.1.7 and B.1.351, respectively. In contrast, healthy HCWs without a prior SARS-CoV-2 infection demonstrated a wide range of nAb titres post-vaccination with responses significantly lower than HCWs with prior SARS-CoV-2 infection. Neutralisation of VOCs with the E484K mutation (B.1.351 and P.1) were consistently lower in HCWs in the absence of evidence of prior SARS-CoV-2 infection (p<0.001). Notably, in vaccinated HCWs with prior SARS-CoV-2 infection, there was a significant increase of neutralising titres post-vaccination to all variants, compared to their pre-vaccination neutralisation titres. This underscores the importance of vaccination to boost neutralising antibody breadth to VOCs, and also provides support for the hypothesis that repeated immunisations will boost protective immunity in individuals without prior SARS-CoV-2 exposure.

## Manuscript

The UK’s population is only partially vaccinated with many having received just one vaccination dose (either BNT162b2 (Pfizer) or AZD1222 (Vaxevria, AstraZeneca)), and tracking the spread of SARS-CoV-2 Variants of Concern (VOCs) remains critical to understanding the levels of vaccine-induced immunity and vaccine-escape variants. The immune correlate of protection to SARS-CoV-2 and COVID-19 established in Phase 3 clinical trials following two doses of vaccine was the level of virus-neutralising antibodies (nAbs) to SARS-CoV-2 in study groups, before the VOCs emerged.^1^ Vaccination programmes are leading to promising reductions in disease severity and mortality in vaccinated populations. However, the combined situation of ongoing transmission within communities with vaccine recipients, and newly arising VOCs, continues to pose a serious threat to public health and the efficacy of these vaccines. Currently in the UK, the interval between the first and second dose of vaccination has been extended beyond 12 weeks. This achieved the aim of maximising population coverage by immunising the greatest possible number of individuals to prevent disease and hospital admissions. Encouragingly, a growing number of studies have reported a marked reduction of cases with moderate-to-severe clinical symptoms and a dramatic decline in hospitalised COVID-19 cases in the UK, underscoring the success of this strategy.^2,3^

Many countries, both in early and advanced stages of vaccine immunisation campaigns, are experiencing new cases of VOCs that have acquired mutations facilitating increased transmission and evasion of pre existing immunity. These VOCs may cause increased morbidity and mortality.^4–6^ Three VOCs, B.1.1.7, B.1.351, and P.1, have been reported in over 114, 68, and 37 countries, respectively, and new cases continue to be reported worldwide. Additional VOCs, such as B.1.617.2, are likely to continue to emerge and threaten current COVID-19 vaccination programmes. Recent reports suggest that both single dose and two-dose vaccine campaigns are exhibiting gaps in protection.^7,8^ A two-dose regimen of the AZD1222 vaccine in South Africa did not show protection against mild-to-moderate COVID-19 accompanying the B.1.351 variant.9 Furthermore, a report of BNT162b2 immunised individuals in Qatar found that its efficacy was 20% lower against B.1.351, compared with B.1 (vaccine strain), with significant breakthrough infections.10 Similar concerns were also noted in a case-cohort study in Israel, which found disproportionately high infection rates with B.1.351 in fully vaccinated compared with unvaccinated individuals.^11^ These findings suggest that despite aggressive immunisation programmes, the presence of circulating VOCs represent a serious concern for the emergence of more virulent vaccine-escape variants.

Understanding the threshold levels of protective immunity against VOCs in specific risk groups and in the population with ongoing transmission is important for informing and refocusing immunisation strategies. This includes targeting specific vulnerable groups, as well as providing data on the immune correlates of protection for the development of effective next generation variant-resistant vaccines. The UK has more than 3.5 million people on the Shielded Patient List which comprises those that are clinically extremely vulnerable and often prone to persistent infections, including cancer and transplant patients. There is a greater risk of higher morbidity in shielding patients, and those with the inability to clear infections, such as immunodeficient patients, may themselves amplify new viral variants.^12^

We undertook a single-centre cross-sectional study of outpatients with immunodeficiencies (IDPs) and health care workers (HCWs) employed at the same critical care tertiary NHS Trust, in order to understand the spectrum of immunity to the vaccine strain compared to three major VOCs following one dose of either BNT162b2 or AZD1222 vaccines. To determine the current level of immunity to VOCs in healthy populations, we compared nAbs induced by either BNT162b2 or AZD1222 in vaccinated HCWs. In addition, we screened 107 outpatients with a range of immunodeficiencies for the presence of SARS-CoV-2 antibodies. These patients included both primary and secondary immunodeficient patients, with disorders including common variable immunodeficiency and specific polysaccharide antibody deficiency syndrome. For both the HCWs and IDPs, we determined their ability to neutralise the B.1 vaccine strain as well as the three VOCs, B.1.1.7, B.1.351, and P.1. We used a validated SARS-CoV-2 pseudovirus neutralisation assay standardised against WHO reference standards (NIBSC) to determine nAb titres.^13^

Our data revealed either undetectable antibodies or low nAbs in IDPs, with only 5% and 3% showing detectable neutralisation of B.1.1.7 and B.1.351, respectively (Figure A). While a number of the vaccinated HCWs without a prior SARS-CoV-2 infection (Figure B) had a range of neutralising titres, they were markedly lower than HCWs with previous SARS-CoV-2 infection (Figure C). Of the HCWs without a prior SARS-CoV-2 infection, less than 50% had detectable nAbs to B.1; <40%, <10% and <10% had nAbs against B.1.1.7, B.1.351 and P.1, respectively. In contrast, with the exception of some poor responders, vaccinated HCWs with prior infection had marked nAb responses (ranging >3,000 to 10,000 IC_50_ values, or >5,000 to >34,000 International Units) against B.1. Notably, in HCWs with prior SARSCoV-2 exposure, there is a significant increase of neutralising titres post-vaccination to all four strains, compared to their pre-vaccination neutralisation titres (Figure D). This underscores the importance of vaccination even in populations that have been previously exposed, and also provides support for the hypothesis that a second-dose of vaccination will similarly boost immunity to greater protective levels in individuals without prior SARS-CoV-2 exposure.

Further, neutralising titres to B.1 were highest in vaccinated HCWs with a history of prior infection, with no statistical difference between the responses induced by BNT162b2 or AZD1222 vaccines (Fig B&C, p>0.05). Notably, neutralisation of VOCs with the E484K mutation (B.1.351 and P.1) were consistently lower in HCWs in the absence of evidence of prior SARS-CoV-2 infection (Fig B, p<0.001). The key finding was the large spectrum of nAb titres, and the overall low levels of nAbs especially to B.1.351 and P.1 in single-dose vaccinated HCWs, particularly in those without a prior SARS-CoV-2 infection. Further, with the exception of one immunodeficient patient with a history of SARS-CoV-2 infection, titres were 5-fold lower in IDPs than HCWs (Fig A), well below levels of nAbs that correlated with protection in Phase 3 clinical trials to viruses circulating early in the pandemic. These data not only underscore the risk of clinically vulnerable patients to infection with SARS-CoV-2, but also of individual HCWs with no or low neutralising antibodies to the VOCs for onward transmission. This emphasises the importance of extended surveillance of the threshold of neutralising antibodies that correlate with protection from infection and/or disease, against circulating VOCs. Larger population studies will be needed to determine the threshold levels of nAbs across representative cohorts of the UK population. This will help determine risk groups for follow-up immunisations with current vaccines, or variant-specific successors.

Virus neutralisation threshold studies will support targeted vaccination in outbreak communities, and the strategic or targeted use of variant-specific vaccines in the highest risk populations. We suggest that this include HCWs with low nAbs titres and clinically vulnerable IDPs. Understanding the existing levels of immunity to the VOCs in the overall population will inform containment strategies in communities at risk of local VOC outbreaks. Further, this will help to define public health strategies to contain and prevent novel variants of interest or the emergence of vaccine resistant variants that could threaten COVID-19 immunisation programmes.

**Figure:**
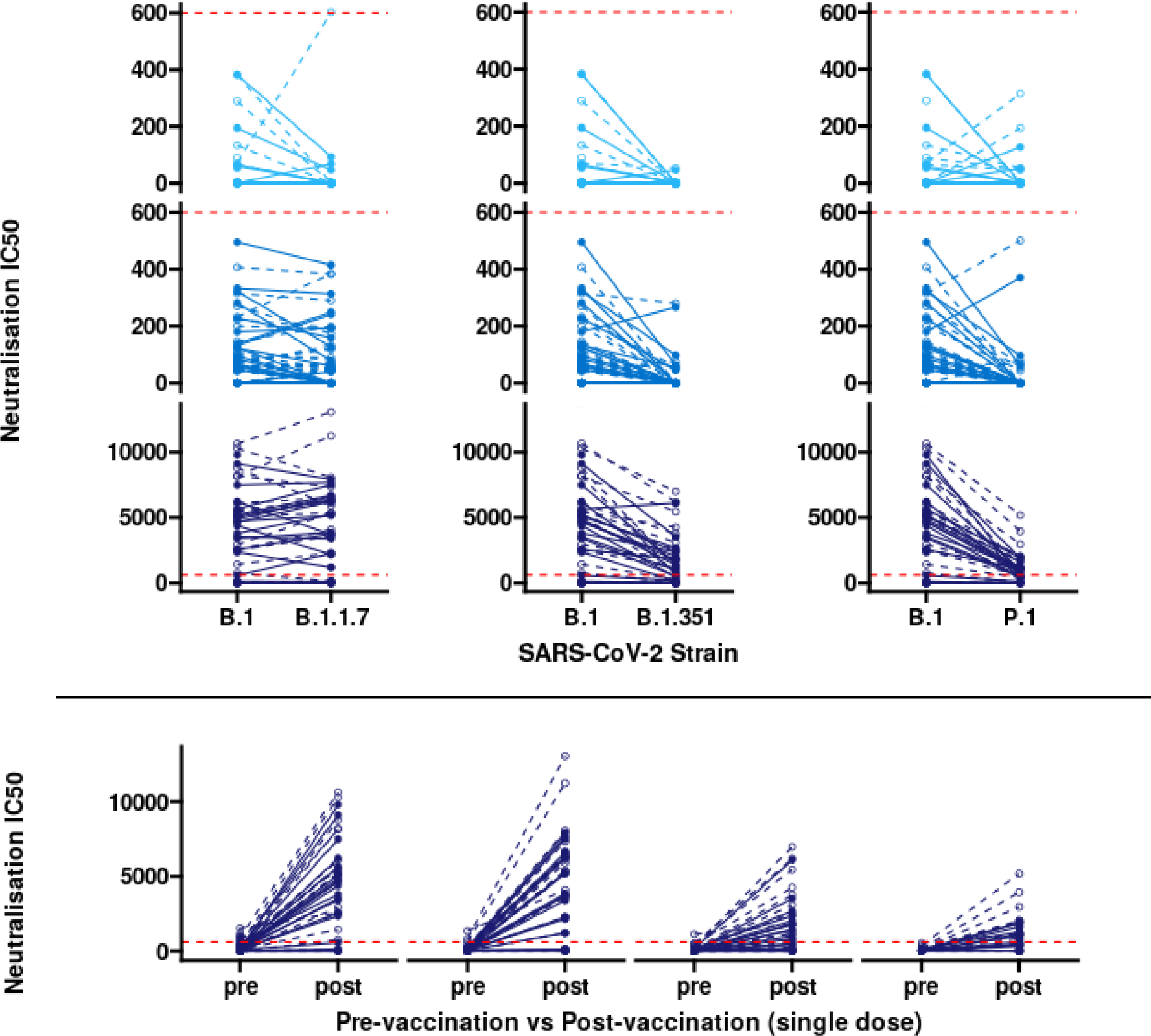

Healthcare workers (HCW) and immunodeficient patients (IDP) immunised with a single-dose of either BNT162b2 (open circle) or AZD1222 (closed circle) demonstrate varied neutralising responses, individuals without prior exposure show lowered responses. All tested groups demonstrate reduced neutralisation against B.1.351 and P.1 relative to the B.1 strain after one dose. A red dashed line indicating the highest IC50 value in plots A and B has been included as a reference to visually compare these low responses with the high responses in C and D.

Fig A, (top row) Immunodeficient patients with both primary and secondary immune deficiencies (n=91) show limited neutralisation (11/91) against the B.1 strain, and overall low neutralising titres (range: negative to IC50 values of 382, geometric mean=1.8). IDPs demonstrate a 1.6-fold reduction in IC50 against B.1.351 (p=0.005), but no significant reduction against B.1.1.7 (1.4-fold, p=0.064). Fig B, (second row) HCWs without SARS-CoV-2 infection prior to immunisation exhibit a large variety of neutralising titres (range: negative to IC50 values of 495, geometric mean=8.1) against B.1, and demonstrate a weakly significant reduction in neutralising titres against B.1.1.7 (1.4-fold reduction, p=0.032). These HCWs show a more substantial 5.3- and 5.1-fold reduction in neutralising titres against B.1.351 and P.1, respectively compared to B.1 (p<0.001). Fig C, (third row) HCWs with SARS-CoV-2 infection prior to immunisation demonstrate increased neutralising titres post-vaccination (range: negative to IC50 values of 10635, geometric mean=1365). As with the other cohorts, these HCWs demonstrate a reduction in neutralising antibody titres against B.1.351 (3.3-fold reduction, p<0.001) and P.1 (4.9-fold reduction, p<0.001), but not B.1.1.7 (p=0.97), compared to the B.1 strain. Despite the reduction in neutralising titres, HCWs with prior infection pre-vaccination demonstrate 19-, 8-, and 10-fold higher levels of neutralising titres against the B.1.1.7, B.1.351 and P.1 compared to HCWs without prior exposure. Fig D, (fourth row) HCWs (from Fig C) with SARS-CoV-2 infection prior to vaccination, demonstrate a significant increase of nAb titres post-single-dose vaccination against all strains (p<0.001)

## Data Availability

All raw data will be made available upon request to corresponding author.

